# Combining longitudinal data from different cohorts to examine the life-course trajectory

**DOI:** 10.1101/2020.11.24.20237669

**Authors:** Rachael A. Hughes, Kate Tilling, Deborah A. Lawlor

## Abstract

Longitudinal data are necessary to reveal changes within the same individual as they age. However, rarely will a single cohort capture data throughout the lifespan. We describe in detail the steps needed to develop life-course trajectories from cohorts that cover different and overlapping periods of life. Such independent studies are likely from heterogenous populations which raises several challenges including: data harmonisation (deriving new harmonised variables from differently measured variables by identifying common elements across all studies); systematically missing data (variables not measured are missing for all participants of a cohort); and model selection with differing age ranges and measurement schedules. We illustrate how to overcome these challenges using an example which examines the effects of parental education, sex, and ethnicity on weight trajectories. Data were from five prospective cohorts (Belarus and four UK regions), spanning from birth to early adulthood during differing calendar periods. Key strengths of our approach include modelling trajectories over wide age ranges, sharing of information across studies and direct comparison of the same parts of the life-course in different geographical regions and time periods. We also introduce a novel approach of imputing individual-level covariates of a multilevel model with a nonlinear growth trajectory and interactions.

---

Life-course epidemiology aims to elucidate biological, behavioural, and psychosocial processes that operate across an individual’s life-course to influence the development of disease risk [1]. It requires repeatedly assessed data on risk factors and trajectories that reflect the underlying propensity of disease risk from infancy to adulthood [1]. Trajectories of underlying disease propensity are necessary to establish ages of ‘peak’ health for different diseases and the extent to which intervening during periods of developmental origins or age-related decline are likely to maximise population health. However, rarely will a single cohort have data on a given risk factor or outcome throughout the lifespan.

An alternative to examining life-course trajectories in a single cohort is to model longitudinal data from multiple cohorts that cover different and overlapping periods of life. Examples include models of longitudinal blood pressure data from eight UK cohorts from ages 7 to 80 [2], alcohol consumption from age 15 years to over 90 years from nine cohort studies [3] and height and weight trajectories from birth to 18 years using sparse longitudinal data from 16 studies [4]. Whilst these studies demonstrate the potential of obtaining life-course trajectories by combining data from several cohorts that cover different periods of the life-course, they do not describe the steps required to overcome the challenges in conducting this type of analysis.

This paper investigates the challenges in combining data from independent cohorts with repeated measurements that cover different and overlapping periods of life. We describe in detail the steps needed to model life-course trajectories from several cohorts (summarised in box 1), including our code in statistical software Stata.

## Box 1. Key challenges

<u>Data harmonisation</u>

- Information may be lost during the data harmonisation process in order to include cohorts with a relatively crude measure of a variable.
- Some variables may not be measured by all cohorts. Subject-matter knowledge may provide information about their most likely values.

<u>Accounting for the data’s dependence structure</u>

- The data’s dependence structure must be appropriately modelled in order to obtain appropriate standard errors.
- Reliable estimation of the random effects’ variance components at a particular level depends on the number of units at that level and not the total number of level-1 observations (e.g., repeated measurements).

<u>Model selection across multiple cohorts</u>

- Model selection on the combined data from all studies may be impractical due to the large volume of data.
- Two alternatives are: (1) fit the same model separately to each cohort and perform model selection on the summed likelihoods across the cohorts. (2) Perform model selection on a random sample of individuals, keeping the proportion of individuals from each cohort as per the combined data.
- Model selection based on summed likelihoods may not be feasible when the feature under comparison (e.g., nonlinear trajectory) requires each cohort to cover the same age range (e.g., restricted cubic splines).

<u>Missing data in the outcome (repeated measurements) and the covariates</u>

- Determining the amount of missing outcome data is difficult for cohorts without a prescribed measurement schedule (such as opportunistic health visits).
- Likelihood estimation of a multilevel model can utilise all available observed outcome data and is valid under the missing at random assumption (i.e., differences between the observed and missing data are explained by associations with the observed outcome and covariate data).
- When outcome data are suspected missing not at random, multiple imputation (MI) can use information from auxiliary variables (not included in the multilevel model) to explain the reasons for the missing data and provide valid inference.
- Restricting the analysis to those with complete covariate data can result in a substantial loss of information especially when covariates are systematically missing in some cohorts (missing for all individuals) and/or when there are many covariates each with small amounts of missing data.
- The imputation model of MI should account for the main features of the multilevel model (e.g., its multilevel structure, any interactions).

## ILLUSTRATIVE EXAMPLE

Our example examines the associations of sex, ethnicity, and parental education on weight trajectories from birth to 20 years. We analysed data from five prospective cohorts: the Avon Longitudinal Study of Parents and Children (ALSPAC) [5, 6], Barry Caerphilly Growth (BCG) study [7, 8], Born in Bradford (BiB) [9], Christ’s Hospital School (CHS) [10], and the PRomotion of Breastfeeding Intervention Trial (PROBIT) [11, 12]. PROBIT is from Belarus and the remaining cohorts are from the UK. Collectively they cover different periods of birth from the 1930s to 2010. Children from multiple births were excluded because their growth patterns differ considerably from singletons. Parental education is a composite variable that takes on all possible joint values of maternal highest educational attainment and paternal occupation (recorded around the time of birth except for CHS). Data were harmonised across the cohorts.

We give a brief description of each cohort study (further details in the Appendix). Table 1 summarises the cohorts’ characteristics.

**Table 1:**
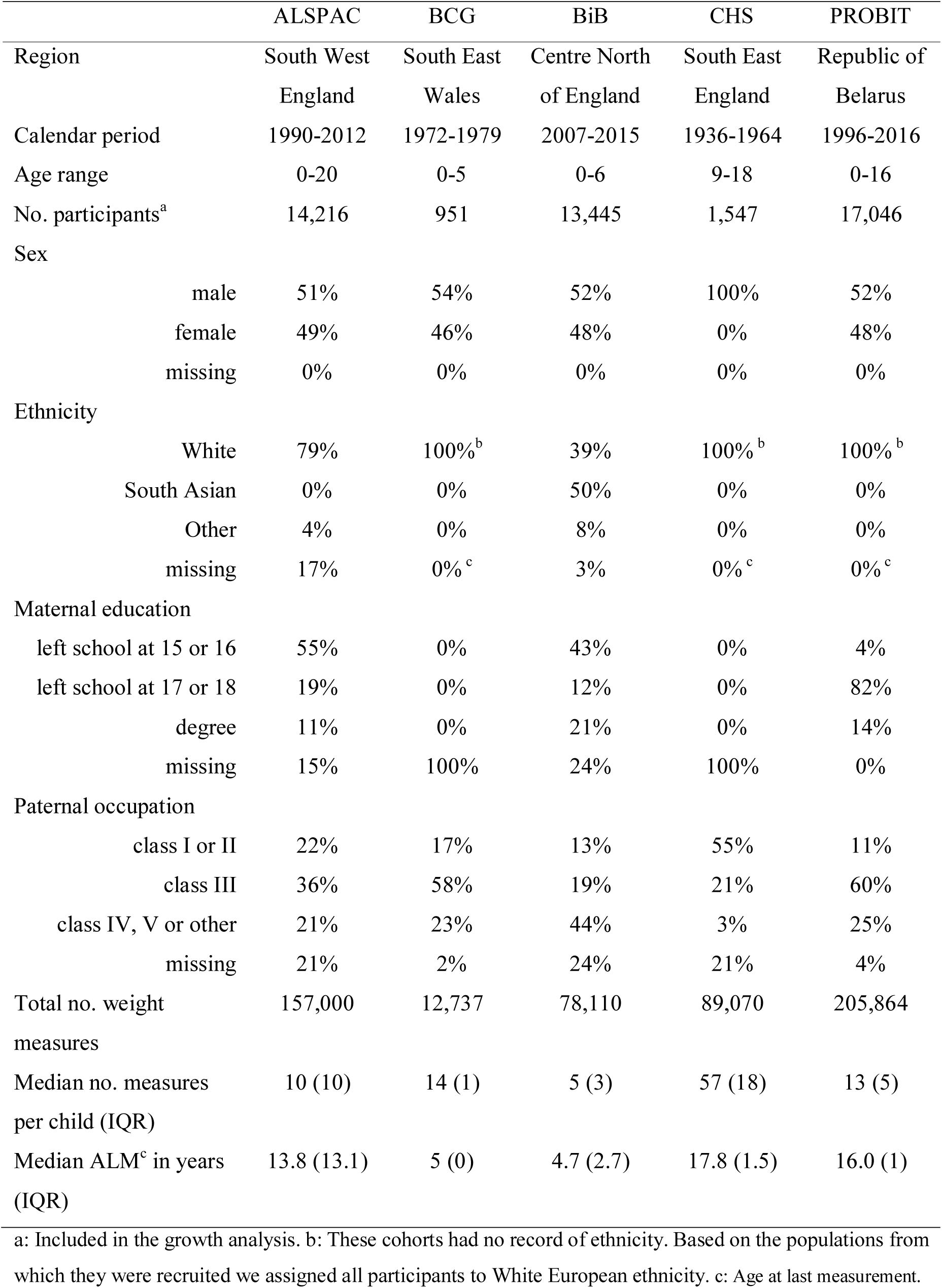
Baseline characteristics of the cohorts using harmonised categories

### Cohort ALSPAC

All pregnant women resident in a defined area in the South West of England, with an expected date of delivery between 1st April 1991 and 31st December 1992, were invited to take part in ALSPAC [5, 6]. The children of these pregnancies have been followed up since birth. Weight measurements were recorded at multiple timepoints between birth and late adolescence (e.g., at birth, 6 weeks, 10, 21 and 48 months, annual measurements between 7 to 11 years, and at target ages 12, 13, 15 and 17 years) [13]. These measurements were from several sources: medical records, research clinics and parent-reports.

### Cohort BCG

BCG is a follow-up of a dietary intervention randomised controlled trial of pregnant women and their offspring [8]. which recruited pregnant women resident in two towns in South Wales between 1972 and 1974 [7]. Birth weights were abstracted from hospital records, and weight measurements were recorded by study nurses who visited at 10 days, 6 weeks, 3, 6, 9 and 12 months, and thereafter at 6-monthly intervals resulting in a total of 14 measurements by age 5 years [8].

### Cohort BiB

All pregnant women booked for delivery at the Bradford Royal Infirmary between March 2007 and November 2010, who attended the oral glucose tolerance test clinic (offered to all women at 26-28 weeks gestation), were invited to take part in the study [9]. The children of these pregnancies have been followed up since birth. Weight measurements were recorded at multiple timepoints between birth and mid-childhood (e.g., birth and taken on average at 2 and 6 weeks, 8 months, 4 and 6 years) [13]. These measurements were from several sources: maternity records, child health records, primary care records, the national child measurement programme (NCMP) and researcher assessments.

### Cohort CHS

Regular measurements were recorded on boys, aged 9 to 18 years, who attended the (all boys) Christ’s Hospital school in West Sussex between 1936 and 1969 [10]. The students were measured three times a school term (i.e. nine measurements per year) by the School medical officer [10].

### Cohort PROBIT

Thirty-one maternity hospitals and associated polyclinics (outpatient clinical for routine healthcare) in the Republic of Belarus participated in this randomised controlled trial. Mother-infant pairs were recruited during their postpartum hospital stay between June 1996 and December 1997 [11, 12]. Birth weight was abstracted from hospital records and weight was measured at scheduled study visits at 1, 2, 3, 6, 9 and 12 months, and at 6.5, 11.5 and 16 years [14]. Also, weight measurements were abstracted from primary care records.

## CHALLENGE: DATA HARMONISATION

Data harmonisation is required when pooling heterogeneous data from different studies [15]. For comparable but differently measured variables, such as ethnicity, data harmonisation derives new ‘harmonised’ variables by identifying common elements across all studies.

### Continuous measurements and sex

The continuous measurements, age and weight, only require conversion to the same units. All four birth cohorts recorded assigned sex at birth. CHS is a cohort from an all-boys school.

### Child ethnicity

For cohorts BCG, CHS and PROBIT, who did not record ethnicity, we assumed all children were of White European origin because there were very few people from minority ethnic backgrounds in these populations during the cohort recruitment periods. We harmonised the BiB variable to categories “South Asian” and “White European” (representing 50% and 39% of the cohort, respectively), and to “other” for the 8% of Black, mixed race and other ethnicities (Appendix-table 1). And, we harmonised the ALSPAC variable to categories “White European” (79% of the cohort) and “other” (for the 4% from a Black African/Caribbean, South Asian, Chinese, mixed and ethnic backgrounds [16]).

### Maternal education

Since qualifications can differ between countries and, between calendar periods within a country (e.g., in 1987 the UK Ordinary-Level qualification was replaced by the General Certificate of Secondary Education), we defined the harmonised variable by the number of years the mother had spent in education (Appendix-table 2). The two older cohorts, BCG and CHS, did not measure maternal education and so all of their values were set to missing.

### Paternal occupation

BCG and CHS classified paternal Occupational Social Class based on the UK registrar general classification [17] (Appendix-table 3). For ALSPAC and BiB, paternal occupation was categorised using the Office for National Statistics’ Standard Occupational Classification (of 1990 and 2000, respectively). Using [18] as a guide, we mapped the ALSPAC and BiB Standard Occupational Classifications to three occupation classes compatible with those of BCG and CHS (professional or managerial; intermediate; routine or unskilled). The occupation categories of PROBIT cut across social classes (e.g., “service worker” includes occupations from professional to routine, and “manual worker” includes skilled and unskilled manual occupations). So, we used paternal occupation along with paternal highest educational attainment to classify PROBIT fathers to “professional or managerial”, “intermediate” or “routine or unskilled”.

## METHODS

### Challenge: accounting for the dependence structure of the data

The data has a 3-level structure with repeated measurements nested within an individual and individuals nested within a cohort.

The data has a nested 3-level structure (repeated measurements nested within an individual and individuals nested within a cohort). There are two main approaches for modelling multilevel data: marginal models (e.g., generalized estimating equations models) and mixed effects models (also known as random effects, multilevel and hierarchical models) [19, 20]. We shall focus on the linear mixed effects model (hereafter referred to as a multilevel model) which can be used to make inferences about changes in the population mean response and to examine the data’s dependence structure (e.g., comparison of within-individual to between-individual variability).

Briefly, a multilevel model consists of “fixed effects” and “random effects” [21]. The fixed effects describe the average relationship between the repeated measurements and time, which we shall call the “average growth trajectory”. The random effects are defined at each level of the data and describe the multiple sources of random variability in the data (e.g., random variability between cohorts, between individuals of the same cohort, and within an individual).

Restricted maximum likelihood estimation is recommended because maximum likelihood estimation of the variance parameters (e.g., variance of a random effect) are biased downwards [21].

In our example, an important consideration was the small number of units at level-3 (i.e., 5 cohorts). Although, there is no clear rule of thumb regarding sample size requirements [22], a simulation study investigating multilevel modelling of units with 1000 observations each showed that at least 25 units was required for reliable inference about the variance parameters [22]. Therefore, we decided to fit a 2-level model (repeated measurements at level-1 and children at level-2) with cohort included as a categorical variable in the fixed effects with interactions between the cohort variable and the trajectory terms to allow each cohort to have its own average growth trajectory.

### Modelling the nonlinear growth trajectory

Most biological growth processes show nonlinear changes over time [13]. Two common approaches to modelling nonlinear growth are fractional polynomials [23] and restricted cubic splines (also known as natural splines) [24].

A natural spline divides the growth trajectory into distinct segments where adjacent segments are joined at knot points. A separate curve is fitted to each segment, where the first and last segments are restricted to be linear, and cubic polynomials for the interior segments.

A fractional polynomial models the entire trajectory using a single, special type of polynomial that can include logarithms (e.g., *time*^0^ = ln *time*), minus powers (e.g., *time*^−2^= 1/*time*^2^), non-integer powers (e.g., 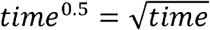) and repeated powers (e.g., *time*^2^, *time*^2^). The degree of the model indicates the number of fractional polynomial terms allowed (e.g., *time*^2^ is a one-degree fractional polynomial, and time^2^, time is a two-degree fractional polynomial).

In our example, we considered natural splines with 3 to 7 knots, where the knots were placed at percentiles (of age) recommended by Harrell [24], and fractional polynomials of one and two degrees with powers −2, −1, 0, 0.5, 1, 2, 3.

### Modelling the covariance structure

The covariance structure of the multilevel model accounts for the dependency within the data and is modelled via the random effects. Misspecification of the covariance structure can lead to incorrect standard errors for the fixed effects and invalid estimates of the random effects’ variance components [21].

At each level, the random effects are assumed to be normally distributed with zero means and an estimated variance-covariance matrix, where the diagonal elements are the variances of the random effects and the off-diagonal elements are pairwise covariances between the random effects. Many software implementations allow the analyst to place restrictions on the variance-covariance matrix (e.g., restrict a covariance to be 0). Typically, the random effects at each level are assumed to be mutually independent of the random effects at all other levels.

Including individual-level random effects for the intercept and slope term(s) allows the growth trajectories (i.e., starting positions and rate of change) to vary between individuals. Other variables, such as sex, can also be included as individual-level random effects.

The measurement-level random effects describe how an individual’s measurements vary about his/her growth trajectory. The simplest structure is a single random intercept which assumes the measurement-level variation is constant throughout. Alternatively, the model can allow the measurement-level variation to depend on age (known as complex level-1 variation [13]).

In our example, all models included individual-level random effects for the intercept and all trajectory terms. We did not put any constraints on the individual-level variance-covariance matrix (i.e., distinct variances and covariances were allowed).

Assuming a constant measurement-level variance was implausible for our data due to the wide age span (i.e., weights at age 18 years were more variable (e.g., 45-75 kg) than birth weights (e.g., 2.0-4.5 kg)). Therefore, we allowed for complex-1 variation, comparing two approaches: (1) included an intercept and linear term for age as measurement-level random effects. (2) Divided the age range into distinct segments and specified a measurement-level random intercept for each segment, where measurement-level variance could vary across the segments but was assumed to be constant within each segment. For both approaches we restricted all pairwise covariances (of the measurement-level variance-covariance matrix) to be zero.

### Challenge: model selection across multiple studies

Applying model selection separately within each cohort can bias selection towards simple functional forms as single studies may lack power to select complex functional forms [25, 26]. Conversely, conducting model selection on the combined data from all studies may be impractical due to the large volume of data.

We considered two alternative approaches: (1) Perform model selection on the summed likelihood across the cohorts (i.e., the same model is fitted separately to each cohort and the likelihoods are summed across the cohort) [27]. (2) Perform model selection on a random sample of individuals from the combined data, where the proportion of individuals from each cohort is the same as per the combined data. An advantage of this second approach is that the remaining data (i.e. the unselected individuals) can be used as a “validation dataset” to further evaluate model fit.

Selection criteria used to compare non-nested models include Akaike Information Criterion (AIC), Bayesian Information Criterion (BIC), and the mean squared prediction error (MSPE) (see Appendix for further information).

In our example, we were not able to compare natural spline modes using the summed likelihood approach due to the cohorts’ differing age ranges (e.g., not possible to fit a model with knot points positioned at the same ages to cohorts with age ranges birth to 5 years, and 9 to 11 years). Instead we selected the top two fractional polynomial and natural spline models based on a random sample of 15,000 children, and then compared the fit of these four models using the validation dataset.

To reduce the number of candidate models, model selection was conducted in two stages. At stage-1 we selected the form of the nonlinear growth trajectory, where all models included the intercept and trajectory terms as fixed effects and individual-level random effects, and the measurement-level random effects were two independent random intercepts for age-periods 0 to 2 years and >2 years. At stage-2 we selected the measurement-level covariance structure of the best-fitting model from stage-1.

### Including covariates

In addition to the cohort variable, we included covariates child’s sex, child’s ethnicity, and parental education as fixed effects and for each covariate we included interactions between the covariate and the trajectory terms. These fixed effects were added to the best-fitting multilevel model.

### Challenge: accounting for missing outcome data

Likelihood estimation of multilevel models utilise all observed repeated measurements and is not biased by the missing data when differences between the observed and missing data can be explained by associations with the observed outcome and covariate data (known as missing at random [28]). In the absence of auxiliary information (i.e., data on variables not included in the multilevel model) this likelihood-based approach makes the same assumption about the missing data as standard implementations of multiple imputation (MI) and will yield standard errors with the same or greater precision [29, 28]. See the Appendix for comments on measuring missing outcome data.

For ALSPAC, the age of last measurement indicates that a sizeable proportion of the cohort exited the study prematurely, which was also reflected in the relatively wide variability in the number of measures per child (Table 1). In comparison PROBIT, which had a similar age range to ALSPAC, had lower levels of dropout and lower variability in frequency of measurement. BCG was originally a trial with a prescribed measurement schedule, which reflects the high levels of participant retention and near-constant number of measures per child. CHS was a school-based, so study participant retention was high. Despite its prescribed measurement schedule, there was sizeable variation in the frequency of measurements (lower and upper quartiles were 49 and 67, respectively). This variability was mainly due to children entering the study (i.e. enrolling at the school) at different ages with 47% of children enrolling before age 11 years, 52% between ages 11 and 12, and 1% aged 13 years or older. Since our example did not have any auxiliary information, we decided not to impute the outcome data.

### Challenge: accounting for missing covariate data

Combining data across cohorts raises the problem of systematically missing covariate data (i.e., covariate is missing for all participants of a cohort because it was not measured [30]). Restricting the analysis to participants with observed data on all covariates, which we shall call “complete covariate analysis” (CCA), and MI are two approaches to account for missing data. The choice of approach will depend on the missing data setting [29]. Specialised MI methods (e.g., [31, 32, 33, 34, 35]) and software (e.g. jomo [36] and Stat-JR [37]) are required when the main analysis is a multilevel model [29, 38].

In our example, there were missing data on the child-level covariates ethnicity, and both components of parental education (Table 1). The discussion below excludes the missing data on child’s ethnicity in BCG, CHS and PROBIT as we assumed all children of these cohorts were of White European origin.

We decided to use MI instead of CCA because: (1) our investigations indicated that the chance of having complete covariate data depended on the observed outcome data; thus, a CCA could be biased by omitting children with missing data [29]. (2) CCA would be extremely wasteful, discarding all data on 17.4% of children: all 2498 children of CHS and BCG, and 5734 children of ALSPAC and BiB. Factors identified as predictors of missingness, and the missing values were included in the imputation model. Since the aim of our example was to illustrate our methodology, we only considered a small number of factors.

Neither jomo nor Stat-JR were suitable for our example because they did not allow the measurement-level variance to vary with age and applying an imputation model that assumed constant measurement-level variance failed to converge. Instead we adapted an imputation procedure developed for imputing level-2 data using summary measures of any level-1 data [33]. Our adaption was an iterative procedure which incorporated summaries of the children’s growth trajectories in the imputation model. It accounted for the interactions of the main analysis because we used the same model to derive these child-specific growth trajectory summaries (see Appendix for further details). We generated 25 imputed datasets and combined the multiple sets of results into a single inference using Rubin’s rules [39].

## RESULTS

The five cohorts contributed data from birth to age 20 years. The combined sample size was 47,205 children with 542,781 weight measurements (Table 1). The cohorts covered overlapping but differing age ranges with repeat weight assessments: ALSPAC 0 to 20 years, BCG 0 to 5 years, BiB 0 to 7 years, CHS 9 to 18 years, and PROBIT 0 to 18 years. CHS had the highest median number of weight measurements per child (at least quadruple that of the other cohorts), and the second highest was BCG, with a median of 14 measures.

Figure 1 shows observed mean growth trajectories among the cohorts. The general trajectory shape was nonlinear: curving over in the first year, curving slightly under between ages 5 and 12 years and starting to plateau around age 17 years. Compared to ALSPAC and PROBIT, CHS had a lower growth trajectory.

**Figure 1:**
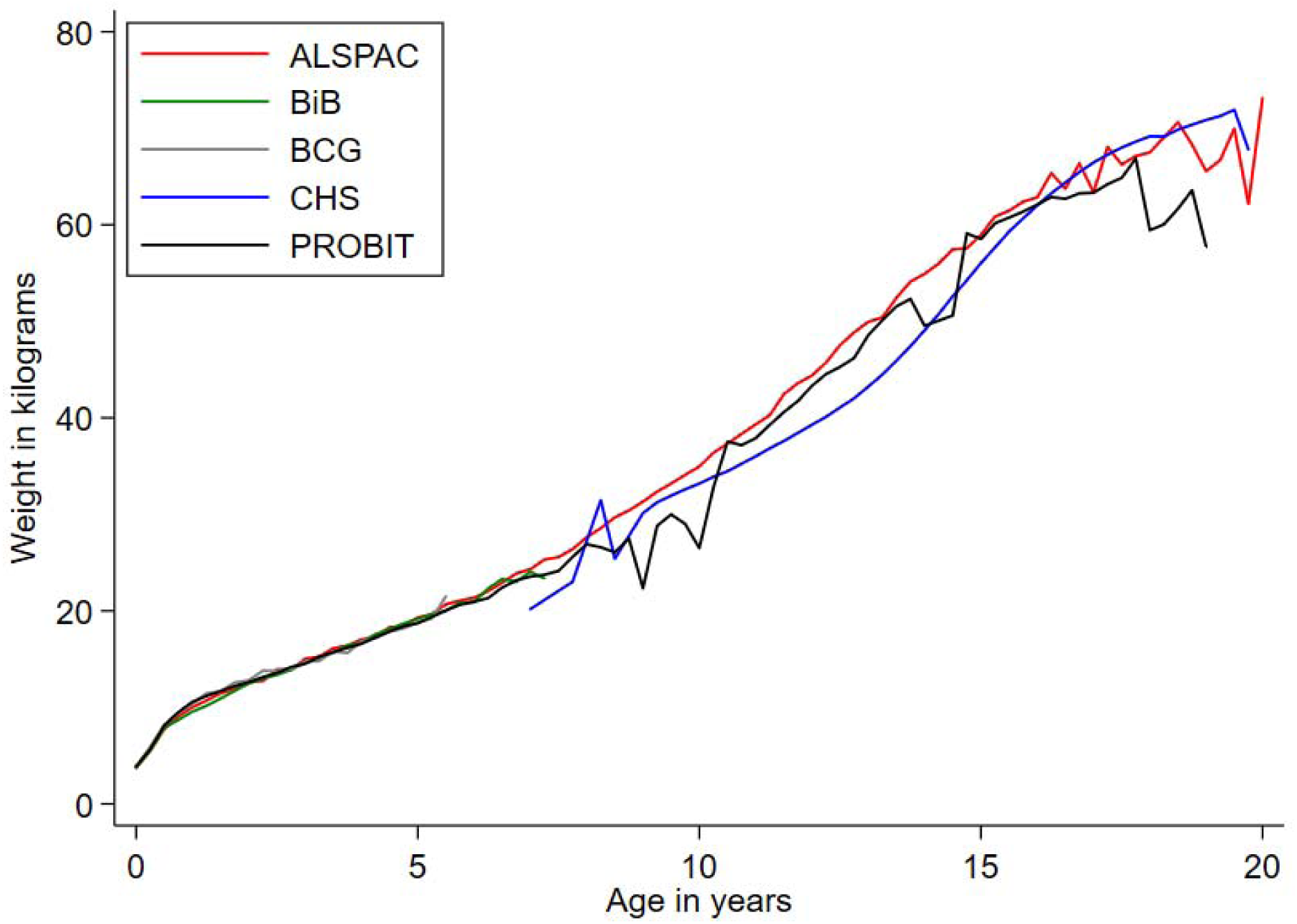
Observed mean weight trajectories among cohorts

The results of the model selection procedure and a summary table of the components of the final model (Appendix-table 7) are reported in the Appendix. Figure 2 shows predicted mean weight trajectories, using the MI point estimates of the final model, between children of different cohorts, sexes, ethnic groups, and parental education groups (in each case holding all other covariates constant). Weight trajectories were very similar between the cohorts in the first four years of life. Between ages 10-15 years children from ALSPAC were heaviest, PROBIT in between and CHS lightest (e.g., at age 15 years the predicted mean difference in weight between children from PROBIT and ALSPAC was −0.88 kg [95% confidence interval (CI) −0.44, −1.33 kg] and between children from CHS and ALSPAC was −6.83 kg [95% CI −6.17, −7.49 kg]). By age 20 years the difference in weight between children from CHS and ALSPAC had narrowed (predicted mean difference −2.84 kg [95% CI −1.57, −4.10 kg]). The marked plateauing effect after age 15 years for PROBIT could be due to its limited number of measures between 15-20 years. The weight trajectories of boys and girls were similar until adolescence and start to diverge after age 15 years. There were very little differences between children of different ethnic backgrounds or between those whose parents had different educational levels.

**Figure 2:**
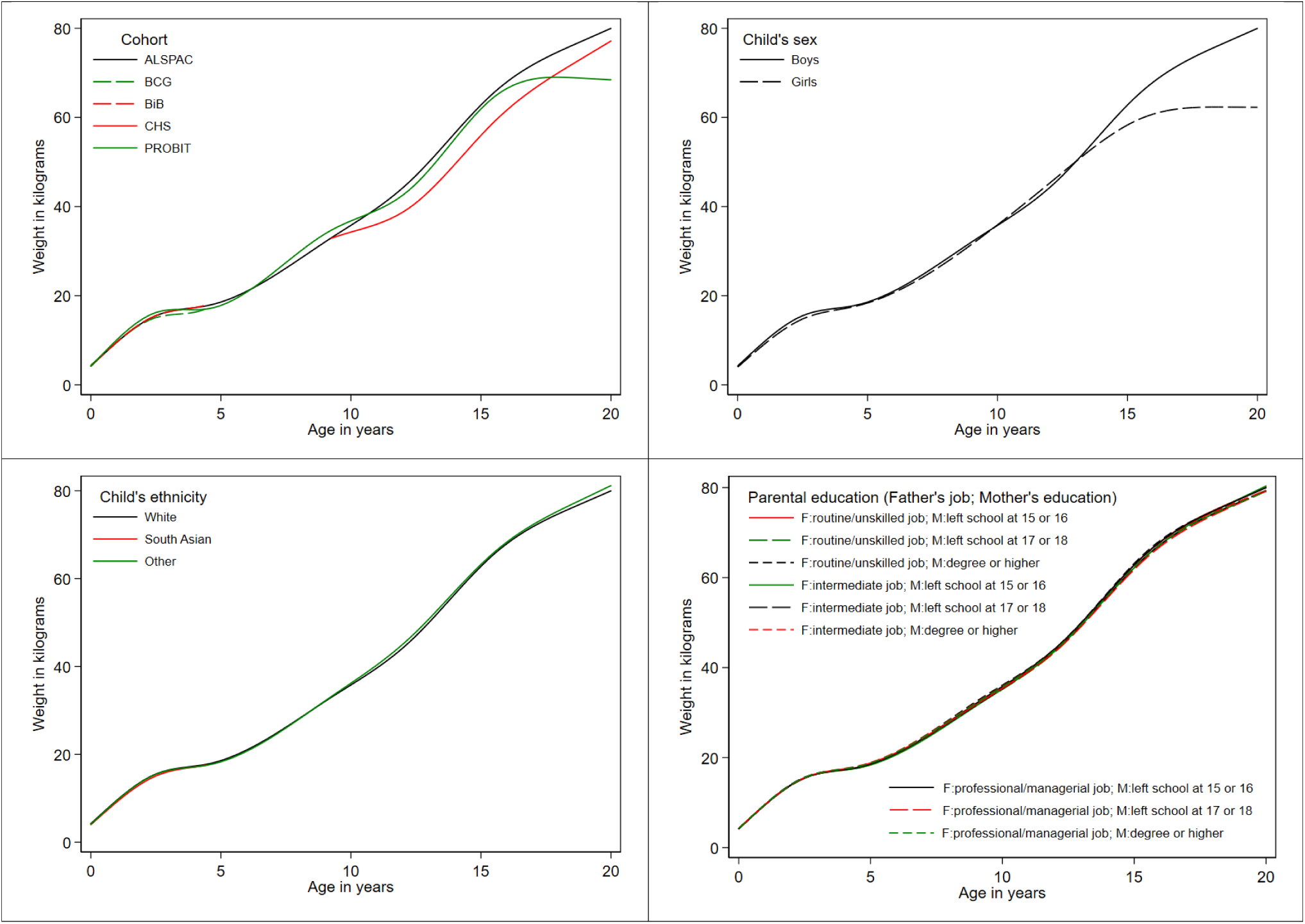
Predicted mean weight trajectories according to (a) cohort study, (b) child’s sex, (c) child’s ethnicity, and (d) parental education. The solid black line in each graph denotes the predicted mean weight trajectory for the reference participant: white boys from the ALSPAC cohort whose mother’s left school aged 15 or 16 from the ALSPAC cohort and father’s occupation is classified as professional or managerial.

## DISCUSSION

Life-course epidemiology should ideally use repeatedly assessed data from when participants were in utero or born through to old age. The oldest birth cohorts are from the 1950s/60s [40, 41], meaning even they only go up to mid-life. These two cohorts also lack very detailed repeat measurements even of weight and height, with substantial gaps between childhood and later adult measures. An alternative is to combine cohorts that cover different but slightly overlapping periods of the life-course to understand the impact of developmental/early life exposures on outcome trajectories across life [2]. We have illustrated the key challenges in this alternative approach and that it not only allows examination of a longer life-course but also direct comparison of the same parts of the life-course in different geographical regions and time periods. We have also introduced a novel approach of imputing individual-level covariates of a multilevel model with a nonlinear growth trajectory and interactions.

In order to cover a life-course it is usually necessary to combine heterogeneous cohorts, in contrast to a meta-analysis of longitudinal studies (e.g., [42]), which selects cohorts for analysis based on their similarity of design. It is also necessary to make some modelling assumptions common to all cohorts in order to gain information over separate modelling of each cohort. This requires the analyst to make decisions on which common assumptions are plausible and which notable differences between the cohorts must be accommodated in the model. In our example, the weight trajectories of each cohort had the same shape and so it was plausible to model all of the data using the same nonlinear trajectory. At the same time, we had to allow for important differences such as a lower mean trajectory for CHS compared to ALSPAC and PROBIT.

Data harmonisation is required when pooling heterogeneous data from different studies. By necessity, the level of detail in the harmonised variable is often determined by the cohort with the most simplified definition of the variable. Also, even after harmonisation the interpretation of the harmonised variable may differ between cohorts (e.g., the meaning of maternal education may change over time or geography).

In meta-analyses, MI has been proposed as a solution to systematically missing covariate data [30, 43, 44] and a study examining early and midlife risk factors for cardiovascular disease in four pooled cohorts imputed exposure variables not available in younger cohorts [45]. In our example we multiply imputed the systematically missing data on maternal education but used a simpler approach for ethnicity since population demographics at the time indicated that almost of all of the participants would have been of White European origin. For cohorts with greater ethnic diversity, external population-level information could be utilised to multiply impute ethnicity as part of a sensitivity analysis [46].

Caution is needed when interpreting parameters common to all cohorts. In our example, the trajectories of boys and girls appeared to diverge from age 15 years onwards. Whilst this may be a real phenomenon we must also consider that this pattern may be unduly influenced by the CHS cohort, which was the only cohort to cover the time-period 1930s to 1950s, included data on boys only and its weight trajectories would likely differ from those of boys from later periods due to changes in lifestyle and increases in the average height.

In summary, careful analysis can harmonise and bring together information from different cohorts to inform life-course trajectories. Our approach can leverage several cohorts to inform about life-course trajectories and examine heterogeneity between cohorts to shed light on influences on trajectories.

## Data Availability

The data are owned by the studies which prohibits me from sharing the data. However, researchers can access the data by submitting requests to the listed studies.

## ACKNOWLEDGEMENTS

We thank the study executives of ALSPAC, BiB, and PROBIT, Christ’s Hospital, and Dr PC Elwood (MRC Epidemiology Unit, South Wales) and Prof. Y Ben-Shlomo (University of Bristol) for permitting access to the data. ALSPAC, BCG study, Born in Bradford, CHS and PROBIT were only possible because of the enthusiasm and commitment of the children and parents. We are grateful to all the participants, health professionals and researchers who have made these studies happen.

## REFERENCES

[1] D. Kuh, Y. Ben-Shlomo, J. Lynch, J. Hallgvist and C. Power, “Life course epidemiology,” J Epidemiol Community Health, vol. 57, no. 10, pp. 778–783, 2003.

[2] A. K. Wills, D. A. Lawlor, F. E. Matthews, A. A. Sayer, E. Bakra, Y. Ben-Shlomo, M. Benzeval, E. Brunner, R. Cooper, M. Kivimaki, D. Kuh, G. Muniz-Terrera and R. Hardy, “Life Course Trajectories of Systolic Blood Pressure Using Longitudinal Data from Eight UK Cohorts,” PLoS Med., vol. 8, no. 6, p. e1000440, 2011.

[3] A. Britton, Y. Ben-Shlomo, M. Benzeval, D. Kuh and S. Bell, “Life course trajectories of alcohol consumption in the United Kingdom using longitudinal data from nine cohorts,” BMC Med., p. 13:47, 2015.

[4] C. Anderson, L. Xiao and W. Checkley, “Using data from multiple studies to develop a child growth correlation matrix,” Stat Med., vol. 38, no. 19, pp. 3540–54, 2019.

[5] A. Fraser, C. Macdonald-Wallis, K. Tilling, A. Boyd, J. Golding, G. Davey Smith, J. Henderson, J. Macleod, L. Molloy, A. Ness, S. Ring, S. Nelson and D. Lawlor, “Cohort Profile: the Avon Longitudinal Study of Parents and Children: ALSPAC mothers cohort,” Int J Epidemiol, vol. 42, no. 1, pp. 97–110, 2013.

[6] A. Boyd, J. Golding, J. Macleod, D. Lawlor, A. Fraser, J. Henderson, L. Molloy, A. Ness, S. Ring and G. Davey Smith, “Cohort Profile: the ‘children of the 90s’--the index offspring of the Avon Longitudinal Study of Parents and Children,” Int J Epidemiol, vol. 42, no. 1, pp. 111–27, 2013.

[7] P. Elwood, T. Haley, S. Hughes, P. Sweetnam, O. Gray and D. Davies, “Child growth (0-5 years), and the effect of entitlement to a milk supplement,” Arch Dis Child, vol. 56, no. 11, pp. 831–5, 1981.

[8] A. McCarthy, R. Hughes, K. Tilling, D. Davies, G. Davey Smith and Y. Ben-Shlomo, “Birth weight; postnatal, infant, and childhood growth; and obesity in young adulthood: evidence from the Barry Caerphilly Growth Study,” Am J Clin Nutr, vol. 86, no. 4, pp. 907–13, 2007.

[9] J. Wright, N. Small, P. Raynor, D. Tuffnell, R. Bhopal, N. Cameron, L. Fairley, D. Lawlor, R. Parslow, E. Petherick, K. Pickett, D. Waiblinger, J. West and Born in Bradford Scientific Collaborators Group, “Cohort Profile: The Born in Bradford multi-ethnic family cohort study,” Int J Epidemiol, vol. 42, no. 4, pp. 978–91, 2013.

[10] J. Sandhu, Y. Ben-Shlomo, T. Cole, J. Holly and G. Davey Smith, “The impact of childhood body mass index on timing of puberty, adult stature and obesity: a follow-up study based on adolescent anthropometry recorded at Christ’s Hospital (1936–1964),” Int J Obes, vol. 30, no. 1, pp. 14–22, 2006.

[11] M. S. Kramer, B. Chalmers, E. D. Hodnett, Z. Sevkoyskaya, I. Dzikovich, S. Shapiro, J.-P. Collet, I. Vanilovich, I. Mezen, T. Ducruet, G. Shishko, V. Zubovich, D. Mknuik, E. Gluchanina, V. Dombrovskiy, A. Ustinovitch, T. Kot, N. Bogdanovich, L. Ovchinikova, E. Helsing and PROBIT Study Group, “Promotion of Breastfeeding Intervention Trial (PROBIT): a randomized trial in the Republic of Belarus,” JAMA, vol. 285, no. 4, pp. 413–20, 2001.

[12] R. Patel, E. Oken, N. Bogdanovich, L. Matush, Z. Sevkoyskaya, B. Chalmers, E. Hodnett, K. Vilchuck, M. S. Kramer and R. M. Martin, “Cohort profile: The promotion of breastfeeding intervention trial (PROBIT),” Int J Epidemiol, vol. 43, no. 3, pp. 679–90, 2014.

[13] L. D. Howe, K. Tilling, A. Matijasevich, E. S. Petherick, A. Cristina Santos, L. Fairley, J. Wright, I. S. Santos, A. J. Barros, R. M. Martin, M. S. Kramer, N. Bogdanovich, L. Matush, H. Barros and D. A. Lawlor, “Linear spline multilevel models for summarising childhood growth trajectories: a guide to their application using examples from five birth cohorts,” Stat Methods Med Res, vol. 25, no. 5, pp. 1854–74, 2016.

[14] R. M. Martin, M. S. Kramer, R. Patel, S. L. Rifas-Shiman, J. Thompson, S. Yang, K. Vilchuck, N. Bogdanovich, M. Hameza, K. Tilling and E. Oken, “Effects of Promoting Long-term, Exclusive Breastfeeding on Adolescent Adiposity, Blood Pressure, and Growth Trajectories. A Secondary Analysis of a Randomized Clinical Trial,” JAMA Pediatr., vol. 171, no. 7, p. e170698, 2017.

[15] B. Rolland, S. Reid, D. Sterling, G. Warnick, M. Thornquist, Z. Feng and J. D. Potter, “Toward Rigorous Data Harmonization in Cancer Epidemiology Research: One Approach,” Am J Epidemiol, vol. 182, no. 12, pp. 1033–8, 2015.

[16] Office for National Statistics, “Bristol 2001 Census Profile,” Bristol.gov.uk, Bristol, 2003.

[17] D. J. Pevalin and D. Rose, “The national statistics socio-economic classification: unifying official and sociological approaches to the conceptulisation and measurement of social class in the United Kingdom,” Sociétés contemporaines, vol. 1, pp. 75–106, 2002.

[18] Office for National Statistics, “The National Statistics Socio-economic class (rebased on SOC2010) user manual,” Palgrave Macmillan, New York, 2010.

[19] G. Fitzmaurice and G. Verbeke, “Parametric modeling of longitudinal data: Introduction and overview,” in Longitudinal Data Analysis, London, Chapman & Hall/CRC, 2008, pp. 31–42.

[20] G. Molenberghs and G. Verbeke, Linear mixed models for longitudinal data, Second ed., New York: Springer-Verlag, 2001.

[21] P. Diggle, P. Heagerty, K. Y. Liang and S. Zeger, Analysis of longitudinal data, Second edition ed., Oxford: Oxford University Press, 2002.

[22] M. L. Bryan and S. P. Jenkins, “Multilevel modelling of country effects: a cautionary tale,” European Sociological Review, vol. 32, no. 1, pp. 3–22, 2016.

[23] P. Royston and D. G. Altman, “Regression Using Fractional Polynomials of Continuous Covariates: Parsimonious Parametric Modelling,” Applied Statistics, vol. 43, no. 3, pp. 429–67, 1994.

[24] F. E. Harrell, Regression Modeling Strategies: With Applications to Linear Models, Logistic Regression, and Survival Analysis., New York: Springer, 2001.

[25] P. Royston and W. Sauerbrei, Multivariable Model-Building: A Pragmatic Approach to Regression Analysis Based On Fractional Polynomials for Modelling Continuous Variables., West Sussex, UK: Wiley, 2008.

[26] W. Sauerbrei and P. Royston, “A new strategy for meta-analysis of continuous covariates in observational studies.,” Statist. Med., vol. 30, no. 28, pp. 3341–3360, 2011.

[27] I. R. White, S. Kaptoge, P. Royston, W. Sauerbrei and The Emerging Risk Factors Collaboration, “Meta-analysis of non-linear exposure-outcome relationships using individual participant data: A comparison of two methods,” Stat Med, vol. 38, no. 3, pp. 326–38, 2019.

[28] J. G. Ibrahim and G. Molenberghs, “Missing data in longitudinal studies: a review,” Test, vol. 18, pp. 1–43, 2009.

[29] R. A. Hughes, J. Heron, J. A. Sterne and K. Tilling, “Accounting for missing data in statistical analyses: multiple imputation is not always the answer,” International Journal of Epidemiology, vol. 48, no. 4, pp. 1294–1304, 2019.

[30] M. Resche-Rigon, I. R. White, J. W. Bartlett, S. A. Peters, S. G. Thompson and PROG-IMT Study Group, “Multiple imputation for handling systematically missing confounders in meta-analysis of individual participant data,” Statistics in Medicine, vol. 32, p. 4890–4905, 2013.

[31] M. H. Huque, M. Moreno-Betancur, M. Quartagno, J. A. Simpson, J. B. Carlin and K. J. Lee, “Multiple imputation methods for handling incomplete longitudinal and clustered data where the target analysis is a linear mixed effects model,” Biometrical Journal, vol. 62, pp. 444–466, 2020.

[32] C. Enders, T. Hayes and H. Du, “A Comparison of Multilevel Imputation Schemes for Random Coefficient Models: Fully Conditional Specification and Joint Model Imputation with Random Covariance Matrices,” Multivariate Behavioral Research, vol. 58, no. 5, pp. 695–713, 2018.

[33] S. Grund, O. Lüdtke and A. Robitzsch, “Multiple imputation of missing data at level 2: a comparison of fully conditional and joint modeling in multilevel designs,” Journal of Educational and Behavioural Statistics, vol. 43, no. 3, pp. 316–353, 2018.

[34] R. Wijesuriya, M. Moreno-Betancur, J. B. Carlin and K. J. Lee, “Evaluation of approaches for multiple imputation of three-level data,” BMC Medical Research Methodology, vol. 20, p. 207, 2020.

[35] C. K. Enders, H. Du and B. T. Keller, “A Model-Based Imputation Procedure for Multilevel Regression Models With Random Coefficients, Interaction Effects, and Nonlinear Terms,” Psychological Methods, vol. 25, no. 1, pp. 88–112, 2020.

[36] M. Quartagno and J. Carpenter, “jomo: A package for multilevel joint modelling multiple imputation,” 2020.

[37] R. Parker and H. Goldstein, “Imputation for multilevel models with missing data using Stat-JR,” Centre for Multilevel Modelling, University of Bristol, Bristol, UK, 2020.

[38] J. R. Carpenter and M. G. Kenward, Multiple Imputation and its Application, Chichester: Wiley, 2013.

[39] D. B. Rubin, Multiple imputation for nonresponse in surveys, New York: John Wiley & Sons, 1987.

[40] P. Rantakallio, “The longitudinal study of the northern Finland birth cohort of 1966,” Paediatr Perinat Epidemiol, vol. 2, no. 1, pp. 59–88, 1988.

[41] D. A. Leon, D. A. Lawlor, H. Clark and S. Macintyre, “Cohort Profile: The Aberdeen Children of the 1950s Study,” Int J Epidemiol, vol. 35, no. 3, pp. 549–552, 2006.

[42] D. O’Neill, A. Britton, M. K. Hannah, M. Goldberg, D. Kuh, K. Tee Khaw and S. Bell, “Association of longitudinal alcohol consumption trajectories with coronary heart disease: a meta-analysis of six cohort studies using individual participant data,” BMC Medicine, vol. 16, no. 124, pp. 1–13, 2018.

[43] S. Jolani, T. P. Debray, H. Koffijberg, S. van Buuren and K. G. Moons, “Imputation of systematically missing predictors in an individual participant data meta-analysis: a generalized approach using MICE,” Statistics in Medicine, vol. 34, pp. 1841–1863, 2015.

[44] M. Quartagno and J. R. Carpenter, “Multiple imputation for IPD meta-analysis: allowing for heterogeneity and studies with missing covariates,” Statistics in Medicine, vol. 35, pp. 2938–2954, 2016.

[45] A. Z. Al Hazzouri, E. Vittinghoff, Y. Zhang, M. J. Pletcher, A. E. Moran, K. Bibbins-Domingo, S. H. Golden and K. Yaffe, “Use of a pooled cohort to impute cardiovascular disease risk factors across the adult life course,” International Journal of Epidemiology, vol. 48, no. 3, pp. 1004–13, 2019.

[46] T. M. Pham, J. R. Carpenter, T. P. Morris, A. M. Wood and I. Petersen, “Population-calibrated multiple imputation for a binary/categorical covariate in categorical regression models,” Statistics in Medicine, vol. 38, pp. 792–808, 2019.

