## Appendix for "Combining longitudinal data from different cohorts to examine the life-course trajectory"

**DESCRIPTION OF THE COHORTS**

Our example examines the associations of sex, ethnicity, and maternal education on weight trajectories from birth to 20 years. We analysed data from five prospective cohorts: ALSPAC [1, 2], BCG study [3, 4], BiB [5], CHS [6], and the PROBIT [7, 8]. Below is a summary description of each cohort study.

**The ALSPAC study**

All pregnant women resident in a defined area in the South West of England, with an expected date of delivery between 1st April 1991 and 31st December 1992, were invited to take part in the study. Of these women there were 14,541 pregnancies which resulted in 14062 live-born children of whom 13988 were alive at age 1 year. At a later date, the study identified children of mothers who were eligible for recruitment but did not respond to the original invitation. Two further recruitment phases (from age 7 years onwards) attempted to recruit all children who would have fitted the original eligibility criteria, whilst excluding those who had previously refused enrolment. In total, there were 14,216 singletons with at least one observed measurement of weight [1, 2]. Ethical approval for the study was obtained from the ALSPAC Ethics and Law Committee and the Local Research Ethics Committees. Informed consent for the use of data collected via questionnaires and clinics was obtained from participants following the recommendations of the ALSPAC Ethics and Law Committee at the time. Until age 16-years the primary caregiver (mostly the child’s mother) gave informed written consent for data collection and its use in research; from age 17-years upwards the child gave their own informed written consent. Please note that the study website contains details of all the data that is available through a fully searchable data dictionary and variable search tool found at <http://www.bristol.ac.uk/alspac/researchers/our-data/>.

The weight measurements were from several sources: medical records, research clinics and parent-reports. Birth weight was extracted from medical records, and routine health visitor measurements were taken on average at six weeks, 10, 21 and 48 months of age.

Additionally, a random 10% of the original cohort have measurements from research clinics between the ages of 4 months and 5 years. From age 7 upwards, all children were invited to annual research clinics. Parent-reported measurements were available across all ages. The combined total number of measurements was 157,000. The median number of measures per child for periods 0 to 5 years, 6 to 12 years, and 13 to 20 years were 5 [interquartile range (IQR) 7-4], 5 [IQR 6-3] and 4 [IQR 4-2], respectively. The data has been described elsewhere along with a detailed analysis of the longitudinal weight measurements [9].

**The BCG study**

The BCG study is a follow-up of a dietary intervention randomised controlled trial of pregnant women and their offspring, who were followed up until age five years. We have used data from the original study which recruited pregnant women resident in two small towns in South Wales (Barry, a seaside town, and Caerphilly, an industrial town) between 1972 and 1974. Of the 1163 pregnant women enrolled in the trial, 951 singleton children completed the trial at age 5 [3, 4].

Birth weights were abstracted from hospital records, and thereafter weight measurements were recorded by study nurses who visited at 10 days, 6 weeks, 3, 6, 9 and 12 months, and thereafter at 6-monthly intervals resulting in a total of 14 measurements by age 5 years. The combined total number of measurements was 12,737. The median number of measures per child between birth and age 5 years was 14 [IQR 14-13]. The data has been described elsewhere along with a detailed analysis of the longitudinal weight measurements [4].

**The BiB study**

All pregnant women booked for delivery at the Bradford Royal Infirmary between March 2007 and November 2010, who attended the oral glucose tolerance test clinic (offered to all women at 26-28 weeks gestation), were invited to take part in the study. Of these women there were 13,776 pregnancies which resulted in 13,740 live-born children and 13,495 singletons with at least one observed measurement of weight. Also, of these 13,495 singletons 1,564 were recruited into a sub-study called BiB1000 [5]. Main caregivers (mostly mothers) have provided informed written consent and ethical approval for all aspects of data collection in BiB was granted by the Bradford National Health Service Research Ethics Committee (ref 06/Q1202/48).

The weight measurements were from several sources: maternity records, child health records, primary care records, the national child measurement programme (NCMP) and from researcher visits. For all children in the cohort, birth weight was abstracted from hospital records, routine health visitor measurements were taken on average at two and six weeks, and eight months and school nurse measurements at ages 4 and 6 years (via linkage with the NCMP). Primary-care measurements were available across all ages. Additionally, researchers made home visits to the children of the BiB1000 sub-study at ages 6,12 and 18 months and 2 and 3 years. The combined total number of measurements was 78,110. The median number of measures per child for periods 0 to 5 years and 6 to 7 years were 5 [IQR 7-4] and 1 [IQR 1-1], respectively. The data has been described elsewhere along with a detailed analysis of the longitudinal weight measurements [9].

**The CHS study**

Regular measurements were recorded on boys, aged 9 to 18 years, who attended the Christ’s Hospital school in West Sussex between 1936 and 1969. These former students were followed-up when aged between 48 and 64 years. Of the 3,175 former students, 1,564 granted permission for researchers to use of their school medical records [6]. The South West Multi-Centre Research Ethics (MREC) approved tracing of this cohort. The approval and support of the data custodians was also received.

The students were measured three times a school term (i.e. nine measurements per year) by the School medical officer. The combined total number of measurements was 89,070. The median number of measures per child for periods 9 to 12 years and 13 to 18 years were 17 [IQR 23-10] and 40 [IQR 47-35], respectively. The data has been described elsewhere along with a detailed analysis of the longitudinal anthropometric measurements [6].

**The PROBIT study**

Thirty-one maternity hospitals and associated polyclinics (outpatient clinical for routine healthcare) in the Republic of Belarus participated in the study. Mother-infant pairs were recruited during their postpartum hospital stay between June 1996 and December 1997. Overall, 17,046 full term singletons, who were being breast-fed, with a birth weight of at least 2,500g and an Apgar score of at least 5 at 5 minutes after delivery were recruited into the study [7, 8]. The institutional review board of the Montreal Children’s Hospital approved all aspects of data collection, and the participating parent/guardian signed informed consent forms in Russian at each phase.

Birth weight was abstracted from hospital records and weight was measured at scheduled study visits at 1, 2, 3, 6, 9 and 12 months, and at 6.5, 11.5 and 16 years [10]. Also, weight measurements were abstracted from primary care records. As part of routine care, the children were regularly measured by their paediatrician between 12 months and 5 years. The combined total number of measurements was 205,864. The median number of measures per child for periods 0 to 5 years, 6 to 12 years, and 13 to 18 years were 10 [interquartile range (IQR) 11-7], 2 [IQR 3-2] and 1 [IQR 1-1], respectively. The data has been described elsewhere along with a detailed analysis of the longitudinal weight measurements [10].

**MODEL SELECTION CRITERIA**

We used two different types of selection criteria to compare our models: information criteria (Akaike Information Criterion (AIC) and the Bayesian Information Criterion (BIC)), and the mean squared prediction error (MSPE). There are several variants of AIC and BIC when applied to multilevel growth models [11, 12]. We used definitions of marginal AIC, marginal AIC for finite samples, and BIC as used by mixed in Stata, lme() in R and SAS Proc Mixed [12]. As the models were fitted using REML estimation, we replaced $N$ (i.e. the total number of observations) with $N-p$ (where $p$ is the number of fixed effect parameters), in the numerator of these formulae, since the restricted likelihood is based on $N-p$ observations [11, 12, 13]. We also considered variants with $N-p$ and $N$ replaced by $M$ (the total number of level 2 units, i.e., children) [11, 14]. See Appendix-table 4 for the formulae of the information criteria we considered. MSPE is the average squared difference between the model predicted measurements (sometimes called “fitted values”) and the observed measurements. To avoid selecting a model that fits extremely well in one age-region of the trajectory but poorly in another age-region, we calculated the MSPE separately in distinct age periods: first year of life, ages 1-2 years, 3-5 years, 6-10 years, 11-15 years, and 16 years and older.

**MISSING DATA**

**Missing repeated measurements**

Calculating the proportion of missing data at each measurement time-point is straightforward for cohorts with a prescribed measurement schedule, such as cohorts BCG and CHS, where all participants were intended to be measured at the same target ages. However, difficulties arise for cohorts, such as ALSPAC, BiB and PROBIT, where measurements were collected from a mixture of scheduled visits, opportunistic health visits (i.e. where the number of and ages of measurement vary between participants) and scheduled visits only intended for a subsample of the cohort. Therefore, alternative indicators are required such as age of last measurement (an indicator of participant retention into the study), and the total number of measurements per individual (an indicator of frequency of measurement which will reflect participant retention, intermittent missingness and the study’s measurement schedule).

**Details of the iterative imputation procedure**

Our adaption included summaries of the children’s growth trajectories in the imputation model. Each child’s growth curve was summarised by his/her own intercept and trajectory terms, which were linear combinations of the fixed effects and random effects (e.g., child’s intercept calculated as reference fixed intercept + random intercept + cohort-intercept interaction + sex-intercept interaction + ethnicity-intercept interaction + parental education-intercept interaction). The interactions of the main analysis allowed the population average growth trajectories to differ between girls and boys, ethnicity groups, cohorts, and categories of parental education. This assumption was accounted for in our adapted procedure because we used the same main analysis model to derive these child-specific growth trajectory summaries. Since the covariates with missing data were used to predict the child-specific growth trajectories, the imputation procedure iterated between generating the child-specific growth trajectories and imputing the child-level covariates using the child-specific growth trajectories (as described immediately below).

Let $Y=\left( Y_{obs},Y_{mis} \right)$ denote our dataset with $Y_{obs}$ and $Y_{mis}$ denoting the observed and missing data, respectively. To start-off the process (i.e., iteration 0) we filled-in the missing values of the child-level covariates using randomly selected observed values. Given the filled-in values from the last iteration $Y^{t-1}=\left( Y_{obs},Y_{mis}^{t-1} \right)$, iteration $t$ of the imputation procedure consisted of the following steps:

1) Fit our multilevel model to the latest filled-in dataset, $Y^{t-1}=\left( Y_{obs},Y_{mis}^{t-1} \right)$, using REML to estimate the parameters of the model. Let $\theta$ denote the parameters of our multilevel model and $\hat{\theta}^{t}$ the latest estimates of $\theta$.
2) To account for the uncertainty from the estimation procedure in step 1, we used a large-sample approximation to draw parameters $\theta$ from the posterior distribution of a linear mixed effects model (i.e., our multilevel model) [15]. Let $\theta^{*t}$ denote the latest draws of $\theta$.
3) We used the latest parameter draws $\theta^{*t}$ to predict the child-specific growth trajectory summaries.
4) We applied standard FCS imputation with the latest child-specific growth trajectory summaries as predictors in the imputation model. FCS was implemented with one cycle to generate a single set of imputed values to give us our latest filled-in dataset, $Y^{t}=\left( Y_{obs},Y_{mis}^{t} \right)$.

We implemented the above procedure with 10 iterations, where the filled-in dataset of step 4 from the 10^th^ iteration forms the imputed dataset. The whole iterative procedure was repeated 25 times to generate 25 imputed datasets fitted our multilevel model separately to each imputed dataset and combined these multiple sets of results into a single inference using Rubin’s rules [16].

Note, we determined the number of iterations required by monitoring the convergence of the imputation procedure as proposed by the ice command in Stata [17]. We ran the imputation procedure for 50 iterations and at the end of each iteration recorded the mean values of the imputed variables (i.e., in our case frequency counts of the categorical variables). For each imputed variable, we generated a trace plot of the frequency counts against the iteration number to determine how many iterations were needed for convergence. Convergence was judged to have occurred when the pattern of the imputed means was random.

Appendix-table 5 describes the regression models used to conduct the standard FCS in step 4 of the iterative procedure above. We did not directly impute parental education but instead imputed its components maternal education and paternal occupation because participants with observed values for paternal education could provide information about missing maternal education values and vice-versa. We omitted variable cohort as predictor from the regression model for maternal education because it was systematically missing in two cohorts. Also, when imputing ethnicity we used predictive mean matching, where imputed values were sampled only from the observed values of ethnicity [18], to avoid imputing impossible values (i.e., South Asian for the ALSPAC cohort).

**RESULTS OF THE MODEL SELECTION PROCEDURE**

**Stage-1: nonlinear growth trajectory**

From the model comparisons based on the random sample of 15,000 children (across the five cohorts), the first and second best-fitting fractional polynomial models were two-degree models with powers 0.5 and 3, and powers 0 and 2, respectively, and the first and second best-fitting natural spline models had 7 and 6 knots, respectively. Appendix-tables 4 and 6 show the information criteria and MSPE values, respectively, for these four models when fitted to the validation dataset (i.e., the data on the remaining 32,205 unselected children). The best fitting model was the natural spline with 7 knots (i.e. the model with the lowest values for all information criteria and MSPE values across all age periods). The MSPE values in the older age ranges indicate that both fractional polynomial models were poorer fits to the data than the natural spline models. Greater differences between the growth trajectories at older ages is also illustrated by the predicted mean growth trajectories shown in Appendix-figure 1.

**Stage-2: measurement-level variance structure**

Modelling the within-child variance using measurement-level random intercepts for distinct age-periods outperformed including age as a linear term in the measurement-level random effects. These distinct age-periods were based on the eight age-periods defined by the seven knot points of the best fitting model from stage 1: 0 to ≤0.25, >0.25 to ≤2.5, >2.5 to ≤4.5, >4.5 to ≤9.25, >9.25 to ≤12.25, >12.25 to ≤15.5, >15.5 to ≤18.75, and >18.75. We fitted a model with eight measurement-level random intercepts (i.e. one for each period) and fitted simpler models by combining across age periods. All AIC values selected a model with seven measurement-level random intercepts, combining age-periods >2.5 to ≤4.5 and >4.5 to ≤9.25. And all BIC values selected a simpler model with six measurement-level random intercepts, combining age-periods >2.5 to ≤4.5 and >4.5 to ≤9.25, and combining age-periods >12.25 to ≤15.5 and >18.75. We decided upon the simpler model with six measurement-level random intercepts, since the differences between the AIC and BIC values of these two models were small, and their MPSE values were very similar.

### **REFERENCES**

| [1] | A. Boyd, J. Golding, J. Macleod, D. Lawlor, A. Fraser, J. Henderson, L. Molloy, A. Ness, S. Ring and G. Davey Smith, "Cohort Profile: the 'children of the 90s'--the index offspring of the Avon Longitudinal Study of Parents and Children," *Int J Epidemiol,* vol. 42, no. 1, pp. 111-27, 2013. |
| --- | --- |
| [2] | A. Fraser, C. Macdonald-Wallis, K. Tilling, A. Boyd, J. Golding, G. Davey Smith, J. Henderson, J. Macleod, L. Molloy, A. Ness, S. Ring, S. Nelson and D. Lawlor, "Cohort Profile: the Avon Longitudinal Study of Parents and Children: ALSPAC mothers cohort," *Int J Epidemiol,* vol. 42, no. 1, pp. 97-110, 2013. |
| [3] | P. Elwood, T. Haley, S. Hughes, P. Sweetnam, O. Gray and D. Davies, "Child growth (0-5 years), and the effect of entitlement to a milk supplement," *Arch Dis Child,* vol. 56, no. 11, pp. 831-5, 1981. |
| [4] | A. McCarthy, R. Hughes, K. Tilling, D. Davies, G. Davey Smith and Y. Ben-Shlomo, "Birth weight; postnatal, infant, and childhood growth; and obesity in young adulthood: evidence from the Barry Caerphilly Growth Study," *Am J Clin Nutr,* vol. 86, no. 4, pp. 907-13, 2007. |
| [5] | J. Wright, N. Small, P. Raynor, D. Tuffnell, R. Bhopal, N. Cameron, L. Fairley, D. Lawlor, R. Parslow, E. Petherick, K. Pickett, D. Waiblinger, J. West and Born in Bradford Scientific Collaborators Group, "Cohort Profile: The Born in Bradford multi-ethnic family cohort study," *Int J Epidemiol,* vol. 42, no. 4, pp. 978-91, 2013. |
| [6] | J. Sandhu, Y. Ben-Shlomo, T. Cole, J. Holly and G. Davey Smith, "The impact of childhood body mass index on timing of puberty, adult stature and obesity: a follow-up study based on adolescent anthropometry recorded at Christ’s Hospital (1936–1964)," *Int J Obes,* vol. 30, no. 1, pp. 14-22, 2006. |
| [7] | M. S. Kramer, B. Chalmers, E. D. Hodnett, Z. Sevkoyskaya, I. Dzikovich, S. Shapiro, J.-P. Collet, I. Vanilovich, I. Mezen, T. Ducruet, G. Shishko, V. Zubovich, D. Mknuik, E. Gluchanina, V. Dombrovskiy, A. Ustinovitch, T. Kot, N. Bogdanovich, L. Ovchinikova, E. Helsing and PROBIT Study Group, "Promotion of Breastfeeding Intervention Trial (PROBIT): a randomized trial in the Republic of Belarus," *JAMA,* vol. 285, no. 4, pp. 413-20, 2001. |
| [8] | R. Patel, E. Oken, N. Bogdanovich, L. Matush, Z. Sevkoyskaya, B. Chalmers, E. Hodnett, K. Vilchuck, M. S. Kramer and R. M. Martin, "Cohort profile: The promotion of breastfeeding intervention trial (PROBIT)," *Int J Epidemiol,* vol. 43, no. 3, pp. 679-90, 2014. |
| [9] | L. D. Howe, K. Tilling, A. Matijasevich, E. S. Petherick, A. Cristina Santos, L. Fairley, J. Wright, I. S. Santos, A. J. Barros, R. M. Martin, M. S. Kramer, N. Bogdanovich, L. Matush, H. Barros and D. A. Lawlor, "Linear spline multilevel models for summarising childhood growth trajectories: a guide to their application using examples from five birth cohorts," *Stat Methods Med Res,* vol. 25, no. 5, pp. 1854-74, 2016. |
| [10] | R. M. Martin, M. S. Kramer, R. Patel, S. L. Rifas-Shiman, J. Thompson, S. Yang, K. Vilchuck, N. Bogdanovich, M. Hameza, K. Tilling and E. Oken, "Effects of Promoting Long-term, Exclusive Breastfeeding on Adolescent Adiposity, Blood Pressure, and Growth Trajectories. A Secondary Analysis of a Randomized Clinical Trial," *JAMA Pediatr.,* vol. 171, no. 7, p. e170698, 2017. |
| [11] | M. J. Gurka, "Selecting the Best Linear Mixed Model Under REML," *The American Statistician,* vol. 60, no. 1, pp. 19-26, 2006. |
| [12] | S. J. Muller and A. H. Welsh, "Model Selection in Linear Mixed Models," *Statistical Science,* vol. 28, no. 2, pp. 135-67, 2013. |
| [13] | E. F. Vonesh and V. M. Chincilli, Linear and nonlinear models for the analysis of repeated measurements, New York: Marcel Dekker, 1997. |
| [14] | R. E. Kass and A. E. Raftery, "Bayes factors," *Journal of the Americam Statistical Association,* vol. 90, pp. 773-795, 1995. |
| [15] | M. Resche-Rigon, I. R. White, J. W. Bartlett, S. A. Peters, S. G. Thompson and PROG-IMT Study Group, "Multiple imputation for handling systematically missing confounders in meta-analysis of individual participant data," *Statistics in Medicine,* vol. 32, p. 4890–4905, 2013. |
| [16] | D. B. Rubin, Multiple imputation for nonresponse in surveys, New York: John Wiley & Sons, 1987. |
| [17] | P. Royston, "Multiple imputation of missing values: update of ice," *The Stata Journal,* vol. 5, no. 4, pp. 527-536, 2005. |
| [18] | I. R. White, P. Royston and A. M. Wood, "Multiple imputation using chained equations: Issues and guidance for practice," *Statistics in Medicine,* vol. 30, no. 4, pp. 377-399, 2011. |

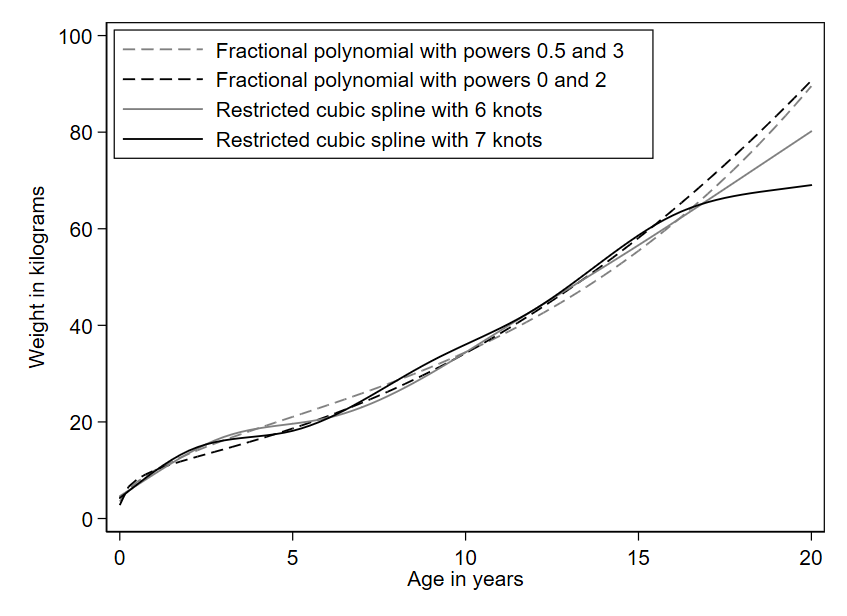

**Appendix-figure 1:** Predicted mean weight trajectories according to the first- and second-best fitting fractional polynomial and natural spline models (generated from fitting the models to all of the combined data).

**Appendix-table 1:** Categories of child’s ethnicity of the harmonised variable and the measured (or assumed) variable in each cohort.

| **Harmonised variable** | **ALSPAC** | **BCG^a^** | **BiB** | **CHS^a^** | **PROBIT^a^** |
| --- | --- | --- | --- | --- | --- |
| White European | White | White British | White British; | White British |  |
|  |  |  | White Other |  | White Other |
| South Asian |  |  | Indian; |  |  |
|  |  |  | Pakistani; |  |  |
|  |  |  | Bangladeshi |  |  |
| Other | Non-White |  | Mixed White and South Asian; |  |  |
|  |  |  | Black; |  |  |
|  |  |  | Mixed White and Black; |  |  |
|  | Unknown |  | Other |  |  |

a: Variable not measured by cohort, assumed category.

**Appendix-table 2:** Categories of maternal education of the harmonised variable and the measured variable in each cohort.

| **Harmonised variable** | **ALSPAC** | **BCG^a^** | **BiB** | **CHS^a^** | **PROBIT** |
| --- | --- | --- | --- | --- | --- |
| Left school at 15 or 16 years | CSE^b^; Vocational qualifications;  O-Level^c^ |  | <5 GCSEs^e^  ≥5 GCSEs |  | Incomplete secondary |
| Left school at 17 or 18 years | A-Level^d^ |  | A-Level |  | Complete secondary;  Advanced secondary;  Partial university |
| Degree or higher | Degree |  | Higher than A-Level |  | Complete university |
| Missing | Missing | Missing |  | Missing | Missing |

a:Variable not measured by cohort, set to missing for all participants. b: Certificate of Secondary Education (CSE). c: Ordinary Level (O-

Level). d: Advanced Level (A-Level). e: General Certificate of Secondary Education (GCSE).

**Appendix-table 3:** Categories of the harmonised variable for paternal occupation and the cohort measured variables of paternal occupation and

paternal highest educational attainment.

| **Harmonised variable** | **ALSPAC**  **(Occupation)** | **BCG^a^** | **BiB**  **(Occupation)** | **CHS^a^** | **PROBIT** | |
| --- | --- | --- | --- | --- | --- | --- |
|  |  |  |  |  | **(Occupation)** | **(Highest education^b^)** |
| Professional or managerial occupation  (Social class I/II) | Professional occupations; Managers and senior officials | Social class I (professional); Social class II (managerial and technical) | Modern professional occupations, senior managers or administrators, traditional professional occupations | Social class I (professional);  Social class II  (managerial and technical) | Service worker | Completed university |
| Intermediate occupation  (Social class III) | Associate professional and technical occupations; Administrative and secretarial occupations; Skilled trades occupations | Social class III (skilled non-manual and manual) | Clerical and intermediate occupations; Technical and craft occupations; Middle or junior managers | Social class III (skilled non-manual and manual) | Service worker | Partial university;  Advanced or complete secondary |
|  |  |  |  |  | Manual worker | Completed or partial university; Advanced or complete secondary |
| Routine or unskilled occupation, other  (Social class IV/V/other) | Personal service occupations;  Sales and customer service occupations; Process, plant, and machine operatives; Elementary occupations | Social class IV (partly skilled); Social class V (unskilled); Student;  Armed forces | Semi-routine manual and service occupations; Routine manual and service occupations; Self-employed; Student; Unemployed | Social class IV (partly skilled); Social class V (unskilled) | Service or manual worker | Incomplete secondary |
|  |  |  |  |  | Farmer, student | All education categories |
| Missing | Missing | Single mother; Unknown; Missing | Unknown | Missing | Service or manual worker; Missing | Missing |
|  |  |  |  |  | Missing | All education categories |

a: UK occupational classification for socio-economic position; b: Highest educational attainment

**Appendix-table 4:** Information criteria from the top two natural spline and fractional polynomial models fitted to the validation dataset

|  | **Formula** | **Natural spline 7 knots** | **Natural spline 6 knots** | **Fractional polynomial**  **powers 0.5 and 3** | **Fractional polynomial**  **powers 0 and 2** |
| --- | --- | --- | --- | --- | --- |
| Marginal Akaike Information Criterion (AIC) | $-2\times{logl}_{R}+2\times s$ | 1403047 | 1506604 | 1500050 | 1510868 |
| Marginal AIC finite sample (using N) | $-2\times{logl}_{R}+\left( \frac{2\times s\times\left( N-p \right)}{N-s-1} \right)$ | 1403047 | 1506604 | 1500050 | 1510868 |
| Marginal AIC finite sample (using M) | $-2\times{logl}_{R}+\left( \frac{2\times s\times M}{M-s-1} \right)$ | 1403048 | 1506604 | 1500050 | 1510868 |
| Bayesian Information Criterion (BIC) (using N) | $-2\times{logl}_{R}+\ln\left( N-p \right)\times s$ | 1403448 | 1506918 | 1500169 | 1510987 |
| BIC (using M) | $-2\times{logl}_{R}+\ln M\times s$ | 1403357 | 1506847 | 1500142 | 1510960 |

Note: logl_R_ is REML-loglikelihood, s is the total number of parameters (i.e. fixed effects and random effects), N is the total number of measurement-level observations, M is the total number of level 2 units (i.e. children), and p is the number of fixed effects parameters.

**Appendix-table 5:** Regression models of the fully conditional specification imputation procedure.

| **Variable to be imputed** | **Regression model** | **Predictors of the regression model** |
| --- | --- | --- |
| Ethnicity^b^ | Multinomial logistic regression | Cohort, maternal education, paternal occupation, sex, child-specific growth trajectory terms^a^ |
| Maternal education | Multinomial logistic regression | Paternal occupation, sex, ethnicity, child-specific growth trajectory terms^a^ |
| Paternal occupation | Multinomial logistic regression | Cohort, maternal education, sex, ethnicity, child-specific growth trajectory terms^a^ |

a: Predicted intercept and trajectory terms for a natural spline with 7 knots. b: Imputed using predictive mean matching.

**Appendix table 6:** Mean squared prediction error in different age periods from the top two selected natural spline and fractional polynomial

models fitted to the validation dataset

| **Age (to nearest year)** | **Natural spline**  **7 knots** | **Natural spline**  **6 knots** | **Fractional polynomial powers 0.5 and 3** | **Fractional polynomial powers 0 and 2** |
| --- | --- | --- | --- | --- |
| Birth to ≤ 1 | 0.61 | 1.09 | 0.24 | 0.37 |
| > 1 to ≤ 2 | 1.38 | 0.90 | 0.95 | 0.56 |
| ≥ 3 to ≤ 5 | 1.25 | 2.30 | 4.86 | 2.20 |
| ≥ 6 to ≤ 10 | 1.11 | 1.44 | 8.91 | 5.34 |
| ≥ 11 to ≤ 15 | 1.75 | 2.35 | 13.13 | 11.80 |
| ≥ 16 | 1.33 | 2.23 | 13.47 | 17.09 |

**Appendix-table 7:** Description of the components of the final multilevel model

| **Model component** | **Description of implementation** |
| --- | --- |
| Fixed effects:  Nonlinear trajectory  Covariates  Interactions |  |
|  | Intercept and six spline terms corresponding to a 7 knot natural spline with knots at 3 months, and 2.5, 4.5, 9.25, 12.25, 15.5 and 18.75 years. |
|  | Categorical variables for cohort, sex, ethnicity, and parental education. |
|  | For each covariate, interactions with the intercept and spline terms. |
| Child-level random effects: | Intercept and spline terms. Assumed to be normally distributed with zeros means and an unstructured covariance matrix (i.e. all variance and covariances are distinct from each other). |
| Measurement-level random: effects | Six random intercepts corresponding to age-periods (in years): 0 to ≤0.25, >0.25 to ≤2.5, >2.5 to ≤9.25, >9.25 to ≤12.25, combined >12.25 to ≤15.5 and >18.75, >15.5 to ≤18.75. Assumed to be normally distributed with zeros means and an independent covariance matrix (i.e. all variances are distinct from each other and zero covariances). |
